# Factors associated with tobacco consumption behavior of adolescent students

**DOI:** 10.1101/2024.05.03.24306835

**Authors:** Anil Kumar Mandal

## Abstract

Although consumption of tobacco is attributed to 8 million deaths of people each year in the world, the consumption of tobacco by adolescent students in Nepal is seen in alarming stage. This paper aims to describe tobacco consumption behavior of adolescent students and its associated factors. For the study, cross-sectional survey design was followed. Students of community schools of Class 10 of Hanumannagar Kankalini Municipality, Saptari were the population of the study. Probability proportional to size method was used to select students. Sample size was 235 but only 225 were included in analysis due to incompleteness of questionnaire of 10 students. Self-administered anonymous questionnaire was used to collect data. Frequency and percentage; and chi-square test were used to analyze descriptive data and inferential data, respectively. Proportion of students who consumed tobacco was 24% (boy-18.7%, girl-5.3%). Sex of students and education of parents were significantly associated with tobacco consumption behavior of students. It is concluded that sex of students, and tobacco consumption behavior of parents are associated factors for high prevalence of tobacco consumption among adolescent students. It should be minimized by promoting girl as role model and educating parents of students.

## Introduction

On not accelerating tobacco cessation program, 8 million deaths will be attributed to tobacco use by 2030 where 80% deaths will occur in low and middle-income countries [1], although, overall prevalence of tobacco consumption among people in the world is in decreasing trend [2]. However, another report has mentioned that 10% of adults death is attributed to tobacco use, and by 2030, tobacco use will take first position to kill people, and 16.67% deaths or 10 million deaths will be attributed to tobacco use among which developing countries will contribute 7 deaths out of 10 [3]. Similarly, another report shows that tobacco use cause eight million deaths in the year in the world [4] that indicates 21,918 people die per day and one people die each four seconds due to tobacco use. While death of 5 million people per year, 13699 people per day and one people each 6.5 seconds was attributed to tobacco use around the decades of 2000 [5]. In the context of Nepal, tobacco kills 27,137 people per year that contribute 14.9% of all deaths in Nepal [6]. In other words, 75 people, 3 people, one people die per year, per hour and per 20 minutes due to tobacco use in Nepal. People who initiates to smoke at their early adolescent stage, as more than 7 in 10 do, and carry it on for 20 years or more will shorten their life 20 to 25 years compared to them who never smoke [5].

Tobacco use is detrimental to health and challenging to economic aspects. In the world, more than 1.4 trillion US dollar is spent for treatment and by losing productivity due to tobacco use [1]. In India, 27.5 billion US dollar was spent for diseases attributed to tobacco use and other aspects in 2017/18, which was 1.04% of total gross domestic product, and only treatment cost of tobacco attributed diseases was 5.3% of total expends of health [7]. A diseased person who consumes tobacco spends 3% of annual income of them on tobacco and its products, and spends 57,215 Nepali rupees in one term of hospitalization, which is higher than average per capita income of Nepali [8].

Although tobacco use has become burden for health and economy, its prevalence is higher especially among adolescent students, which is period of life from 10 to 19 years [9]. They enjoy risk-taking behavior such as tobacco use behavior [10]. As a result, more adolescents of low and middle-income countries of the world and South-East Asian Region of the WHO consume tobacco [2]. In world, average, around 12% or 43.8 million adolescents of 12 to 15 years use at least one or more types of tobacco products. The South-East Asian countries contribute the highest number of adolescents (13-15 years) who consume tobacco. The statistics of it is 43.8 million or 34% of the world’s population of this age group, among them 16% and 8% are boys and girls, respectively. In the low and middle income countries tobacco consumption rate is higher (11-13%) among adolescents of age group 13 to 15 years old than the adolescents of same age in the high-income countries which is less than 10%.

In Nepal, 20.4% adolescents consume any types of tobacco among which 24.6% are boys and 16.4% are girls [11]. Similarly, 9% consume the smoked tobacco among which 11.4% are boys and 6.5% are girls, and 16.2% consume smokeless tobacco among which 19.7% are boys and 12.9% are girls. Similarly, a survey study conducted by Aryal, Bista [12] in correspondence with national health research council found that 9% of adolescent students were current tobacco user among them 11.8% were male and 5.4% were female. It indicates that Nepal is facing burden of tobacco because, for a state, exceeding one million of SLT users or prevalence equal to or more than 10% of SLT consumption is a situation of high burden of SLT for that state [13], for such condition many factors are responsible.

Sex, age, and religion of students, education, occupation, and tobacco consumption behavior of parents; economic status of family are important factors that are associated with tobacco consumption behavior of students. Tobacco consumption behavior of adolescents is highly influenced by tobacco consumption behavior of their parents [14, 15] and other family member at their home and public places [14]. Similarly, tobacco use is higher among men, people with no education or lower level of education, people from lower economic status, and higher age group [16]. Not only this, tobacco consumption has become accepted behavior and a means of meeting with friends and relatives in Nepal [17], and consumption of tobacco among adolescent students [14, 18] is higher in districts of Madhesh Province. In this regard, this paper aims to describe tobacco consumption behavior of adolescent students, and factors associated with it.

## Methods

### Research design and research setting

A cross-sectional survey research design, a non-experimental research where questionnaire or interview is the means of data collection to know the features of population through sample [19], was applied for the study. Community secondary schools of Hanumannagar Kankalini Municipality, Saptari were the study area. Although it is classified as municipality by Government of Nepal, it is mainly a rural area.

### Study variables

Tobacco consumption behavior of students was dependent variable. Sex, age, religion, and place of resident of students; and age, education, occupation, tobacco consumption behavior of parents of students; and economic status of family were independent variables in this paper.

### Measurement of variables

Tobacco consumption behavior of students was identified by asking whether they ever consume any tobacco or not, and parental tobacco consumption behavior was measured in “yes” and “no” by asking whether their parent consume tobacco or not. Sex of students was measured in “male” and “female”. Religion of students was categorized as “Hindu” and “Islam”, whereas place of resident of students was measured as “rural” and “urban” area. Age of students and their parents were measured in continuous form, but, age of students was categorized into “up to 14 years” and “above 14 years”. Likewise, age of parents of students was categorized into “up to 40 years” and “above 40 years”. Education of parents was classified as “no formal education”, and “formal education”. For mother, occupation of mother was categorized into “house work”, “agriculture” and “others”, while occupation of father was categorized into “agriculture”, “business”, “foreign employment” and “others”.

### Population of the study

According to the records of five community schools of Hanumanagar Kankalini Municipality, Saptari, 448 students of Class 10 were population of the study.

### Sample size and sampling

Yamane formula [20] was used to determine sample size at the confidence level was kept 95% and proportion, precision level, and non-response rate were taken 0.5, 5%, and 10%, respectively. On calculation, sample size was 232. To select identified number of sample, proportional stratified sampling procedure was applied. Boys and girls were selected separately in proportion to their enrollment in schools. Although 235 students participated in the study, analysis was performed for 225 students because 10 incomplete questionnaire were not included in the analysis.

### Inclusion and exclusion criteria of participants

Students who were permanent residential of Hanumannagr Kankalini Municipality, Saptari, Nepal and were reading in community secondary school of this municipality were included in the study. Whereas, students who had any mental or health problem, who got married, who were above 19 years old and who did not confirm assent paper were excluded from the study.

### Tools of data collection

Pretested and validated questionnaire was used to collect data that contains questions regarding tobacco consumption behavior of students and relevant demographic aspects of them and their parents. The questions were mainly closed type, and a few are open type questions.

### Data collection technique

The study was conducted from 1 July 2022 to 6 July 2022. Students themselves filled in the anonymous questionnaire in coordination and cooperation of enumerators whom they did not know before the study. Data were collected after providing necessary information to the students in absence of school teacher, however, separate classroom was managed for data collection in coordination of school teacher.

### Data management and statistical analysis

After editing and coding, data were entered into SPSS version 20 for the further analysis. For tobacco consumption behavior of students and parents, and demographic aspects frequency and percentage were used. Chi-square test was applied to identify to what extent tobacco consumed by students associate with demographic aspects of them and their parents.

## Results

In this section, results are presented based on the data analysis procedure in different relevant aspects.

### Demographic information of students

Table 1 shows that over half (54.2%) of the participants were girl, and above half (56.9%) of all respondents belonged age group over 15 years. All most all of them belonged to rural area (89.8%) and Hindu religion (96.4%).

**Table 1.** Demographic information of students, n = 225.

| Description |  | n | % |
| --- | --- | --- | --- |
| Sex | Male | 103 | 45.8 |
|  | female | 122 | 54.2 |
| Age | Up to 15 years | 97 | 43.1 |
|  | 16 years and above | 128 | 56.9 |
| Address | Rural | 202 | 89.8 |
|  | Urban | 23 | 10.2 |
| Religion | Hindu | 217 | 96.4 |
|  | Islam | 8 | 3.6 |

### Demographic information of parents of students

Table 2 provides information about demographic information of parents of students. Majority of students’ father (67.7%) and mother (60%) belonged to age group over 40 years and age group up to 40 years, respectively. Almost all students’ father (83.5%) had achieved formal education whereas just over half of the students’ mother (52.9%) had achieved formal education. Main occupation of students’ father in the study area was agriculture (49.8%) followed by others (28.8%), business (11.4%) and foreign employment (10%). Similarly, main occupation of students’ mother was house work (72%) followed by agriculture (19.1%). Economic status of most of the students’ family was middle class (66.2%) whereas 25.3% and 8.4% students classified their economic status of family as poor and rich, respectively.

**Table 2.** Demographic information of students’ parents, n = 225.

| Description |  | n | % |
| --- | --- | --- | --- |
| Age of father | Up to 40 years | 72 | 32.3 |
|  | 41 years and above | 151 | 67.7 |
| Age of mother | Up to 40 years | 135 | 60.0 |
|  | 41 years and above | 90 | 40.0 |
| Education of father | Informal Education | 37 | 16.5 |
|  | Formal Education | 187 | 83.5 |
| Education of mother | Informal Education | 106 | 47.1 |
|  | Formal Education | 119 | 52.9 |
| Occupation of father | Agriculture | 109 | 49.8 |
|  | Business | 25 | 11.4 |
|  | Foreign Employment | 22 | 10.0 |
|  | Others | 63 | 28.8 |
| Occupation of mother | Housework | 162 | 72.0 |
|  | Agriculture | 43 | 19.1 |
|  | Others | 20 | 8.9 |
| Economic status of family | Poor | 57 | 25.3 |
|  | Medium | 149 | 66.2 |
|  | Rich | 19 | 8.4 |

### Tobacco consumption behavior of students and their parents

Table 3 explicates that among all students, 24% students consumed tobacco, and among them 18.7% were boys and 5.3% were girls. Among boys only, 40.8% boys and among girls only, 9.8% girls consumed tobacco. Supari, paan, gutkha, cigarette, and paan

**Table 3.** Tobacco consumption behavior of students and parents, n = 225.

| Descriptions | n | % |
| --- | --- | --- |
| Students who consumed tobacco | 54 | 24 |
| Students who did not consume tobacco | 171 | 76 |
| Boys who consumed tobacco (among all students) | 42 | 18.7 |
| Boys who consumed tobacco (among all students) | 12 | 5.3 |
| Boys who consumed tobacco (only among boys) | 42 | 40.8 |
| Girls who consumed tobacco (only among girls) | 12 | 9.8 |
| Types of tobacco consumed by students |  |  |
| Gutkha | 18 | 33.3 |
| Khaini | 2 | 3.7 |
| Paan Parag | 9 | 16.7 |
| Paan | 21 | 38.9 |
| Supari | 41 | 75.9 |
| Cigarette | 14 | 25.9 |
| Others | 2 | 3.7 |
| Parents of students who consumed tobacco | 144 | 64 |
| Parents of students who did not consume tobacco | 81 | 36 |

parag were consumed by 75.9%, 38.9%, 33.33%, 25.9%, and 16.7% students, respectively. Likewise, 64% parents of students consumed tobacco.

### Association of tobacco consumption behavior of students with demographic aspects

Table 4 mirrors that sex of students had moderate significant association with their tobacco consumption behavior. Modest significant association was found between tobacco

**Table 4.** Association of tobacco consumption behavior of students with different aspects.

| Descriptions | X<sup>2</sup> | df | *p* | phi |
| --- | --- | --- | --- | --- |
| Sex of students | 29.312 | 1 | .000 | 0.361 |
| Age of students | 1.82 | 1 | 0.177 |  |
| Religion of students | 0.005 | 1 | 0.946 |  |
| Place of resident | 3.29 | 1 | 0.07 |  |
| Age of father | 0.001 | 1 | 0.97 |  |
| Age of mother | 2.961 | 1 | 0.085 |  |
| Education of father | 17.981 | 1 | .000 | 0.283 |
| Education of mother | 5.589 | 1 | 0.018 | 0.158 |
| Occupation of father | 17.386 | 3 | 0.001 | 0.282 |
| Occupation of mother | 5.061 | 2 | 0.08 |  |
| Tobacco consumption behavior of parents | 11.527 | 1 | 0.001 | 0.226 |

*Note.* For non-significant result, phi value is not applicable.

consumption behavior of students and education of both father and mother of students. Tobacco consumption behavior was significantly associated with occupation of father of students and tobacco consumption behavior of their parents with modest effect size.

## Discussion

This paper aims to describe tobacco consumption behavior of students and to find out association between different demographic aspects of students, their parents behavior, and tobacco consumption behavior of students. The study achieved 100% response rate from selected schools and students, and information of 225 students were analyzed to achieve results and conclusion. Higher number of girls participated in the study, and higher proportion of students were above 15 years old. Residential and religion of almost all students was village and Hindu, respectively.

Unlike mother of students, the majority of father of students were from age group above 40 years. Almost all students’ father had access to formal education, however, just over half of the students’ mother had access to formal education that shows the gap of access of education among men and women. Agriculture and house work were major occupation of father and mother of students, respectively. The majority of students categorized their economic status of family as medium.

Percentage of people having no formal education is lower in the report of Nepal Demographic Health Survey (NDHS) 2016 than findings of this study. NDHS 2016 has mentioned that one in ten (10%) men and one-third women have no formal education [21]. Similarly, the report has mentioned that only one-third men is involved in agriculture, which is lower than results of this study. Likewise, over two-third women are engaged in agriculture occupation that is much higher than findings of this study.

One-fourth students of this study ranked economic status of their family as poor that is higher than population of Nepal who are lying below poverty line [22] where it is mentioned that 18.7% population fall under poverty line. However, 31% of people of rural areas belong to the poorest quintiles [21]. The inconsistency among the findings are due to standards taken to identify poverty.

My study shows that more percentage of boys consumed tobacco. Boys consumed tobacco more than three folds compare to girls. Among boys only, above one-third boys consumed tobacco. However, tenth of girls consumed tobacco among girls only. Bhaskar, Sah [23] found 25.3% students consumed tobacco that is slightly higher than findings of my study. But, they reported that 31% boys only among boys and 14.4% girls only among girls consumed tobacco. For boys it is lower and for girls it is higher than the findings of this study. Upreti [18] found 22.8% adolescent students consumed tobacco, which is slightly lower than the findings of this study. Prevalence of tobacco consumption among students was 15.6 % [14] and 19.7% [24] that is lower than findings of this study. Similarly, they found lower proportion of boys and girls who consumed tobacco among boys only and among girls only, respectively. Pradhan, Niraula [24] reported 33.6% boys and 4% girls consumed tobacco, and Chaudhary and Bhandari [14] found that 29.1% boys and 1.3% girls consumed tobacco. Higher proportion of students who consumed tobacco in this might be due to all most all students were from rural areas because prevalence of tobacco consumption among students was higher who were from rural areas (23.5%) than among students who were from urban areas (21.9%) [25].

Consumption of supari was popular among more than three-fourth of students followed by Paan (more than one-third), and Gutkha (one-third). Likewise, cigarette was popular among a quarter of students. Studies vary in their findings. Joshi, Pradhan [26] found that prevalence of consumption of smokeless tobacco and betel nut among students was 6% and 40.5%, respectively. Unlike this study, cigarette was popular among more students (44.4%) than paan parag (25%) and gutkha (11.2%) [18]. Similarly, Chaudhary and Bhandari [14] found that more than two-third (70.8%) students smoked and others consumed smokeless tobacco. However, Bhaskar, Sah [23] reported that only 7.7% students smoked and 17.6% students consumed smokeless tobacco whereas Pradhan, Niraula [24] found that among tobacco user, 17.9% and 8% students ever consumed cigarette and smokeless tobacco, respectively.

This study found that two-third students’ parent consumed tobacco, which is much higher prevalence of tobacco consumption. But, report of NDHS 2016 shows that lower percentage of men (27%) and women (6%) consumed tobacco [21] than what my study found. This difference might be due to different age group of people.

This study found that consumption of tobacco among boys and girls significantly differed. Percentage of tobacco consumer boy significantly differ from percentage of tobacco consumer girls. Study of Chaudhary and Bhandari [14] partially supports our findings. They reported significant association between sex and age of student and tobacco consumption behavior of them whereas religion of students did not significantly associate with tobacco consumption behavior of them. However, sex and religion of students had significant association with tobacco consumption behavior of them [23]. They found that being boy and being from Muslim religion probability of tobacco consumption significantly increase more than two fold (OR = 2.65) and fourfold (OR = 3.90) among students who were boys and who were from Islam religion, respectively.

Education of parents significantly associated with tobacco consumption behavior of students. Prevalence of tobacco consumption was significantly lower among students having parents with formal education than among students having parents with no formal education. Similarly, proportion of tobacco consumption among students was significantly higher among students whose father was involved in agriculture occupation than among students whose father was involved in occupation other than agriculture. Unlike this study, probability of consumption of tobacco among students was higher (OR=1.69) whose father occupation was foreign employment than among students whose father did not went to foreign country for employment [24].

This study resulted that tobacco consumption behavior of parents significantly associated with higher proportion of students who consumed tobacco that is supported by many studies. Having parents [14, 27, 28] and relatives [29] who consume tobacco significantly influence students to consume tobacco. Similarly, tobacco consumption behavior of family member significantly increased likelihood of tobacco consumption among students 11 folds compared to students whose family member did not consume tobacco [23].

This study was based on cross-sectional design and data was collected by using self-administered questionnaire, there might be probability of over or under estimated or both data. Questions regarding generalization might arise as this study included students of only community schools of only one municipality. This study was only limited to tobacco products such as cigarette, Khaini, Gutkha, and Paan Parag including Pan and Supari as these products are mainly consumed with tobacco. Besides the limitations, students were appreciated to provide fact information according to their experience to maintain quality of data.

## Conclusion

My study reveals that prevalence of tobacco consumption among adolescent students is high particularly among boys. Supari, Paan, and Gutkha are common among students. Sex of students, occupation of father, and education and tobacco consumption behavior of parents are significantly associated with tobacco consumption behavior of students. To minimize tobacco consumption among students, girls should be promoted as role model in school and conscious level of parents should be increased through health education intervention.

## Data Availability

The data presented in this study will be available on reasonable request to author.

## Acknowledgements

I would like to hearty thank to the team of Hanumannagar Municiplity, Saptari especially Mr. Surendra Kumar Yadav, former chief executive officer, for their coordination in municipality. Similarly, my thanks goes to school personnel for their coordination, parents of students for their consent, and students who actively participated in the study. I remember enumerators who collected data and coordinated for the study. At last but not least, I am acknowledged to Mr. Chun Chun Yadav who cooperated me during study and edited the language of manusript, and Mr. Bhupendra Prasad Yadav who cooperated me during data collection procedures in the school.

## References

1. National Health Education Information and Communication Centre. FCTC 2030 strategy: Nepal. Kathamandu; 2018.

2. World Health Organization. WHO global report on trends in prevalence of tobacco use 2000-2025. Geneva; 2019.

3. Government of Nepal Ministry of Health and Population. The national anti-tobacco communication campaign strategy for Nepal. Kathmandu; n.d.

4. World Health Organization. WHO report on the global tobacco epidemic 2019: Offer help to quit tobacco use. Geneva; 2019.

5. World Health Organization. The tobacco health toll. 2005.

6. World Health Organization. The fatal link between tobacco and CVD in the WHO South-East Asia Region 2018.

7. John RM, Sinha P, Munish VG, Tullu FT. Economic costs of diseases and deaths attributable to tobacco use in India, 2017-2018 [Accepted Manuscript]. Nicotine and Tobacco Research. 2021;23(2):294–301.

8. Sagtani RA, Pokharel PK, Gurung GN, Pokharel PK. Financial burden of tobacco and catastrophic health care expenditure among smokers admitted in a tertiary care center in eastern Nepal. Nepal Medical College Journal. 2015;17(3-4):125–31.

9. Family Health Division. National Adolescent Health Strategy. 2000.

10. American Psychological Association. Developing adolescents: A reference for professionals. 2002.

11. Nepal 2011. Nepal 2011 (Ages 13-15)Global youth tobacco survey (GYTS) fact sheet. n.d.

12. Aryal KK, Bista B, Dhimal M, Khadka BB, Pandey AR, Mehta R, et al. Global School Based Student Health Survey Nepal, 2015. 2017.

13. Mehrotra R, Sinha DN, Szilagyi T. Global smokeless tobacco control policies and their implementation. Noida -201301, Uttar Pradesh, India; 2017.

14. Chaudhary AK, Bhandari TR. Prevalence of tobacco use and its associated factors among schoolgoing students in Birganj Sub-Metropolitan Nepal. Journal of Health Allied Science 2016;5(1):72–6.

15. Veeranki SP. Advancing Global Tobacco Control: Exploring Worldwide Youth Attitudes and Behaviors toward Tobacco Use and Control [Dr.Ph.]. Ann Arbor: East Tennessee State University; 2012.

16. Nepal Development Research Institute. Taxing tobacco in Nepal: What can be done. 2020.

17. Bajracharya B, Khadka BB, Thapa B. Review of tobacco products used in Nepal especially giving emphasis on smokeless tobacco products. Kathmandu; 2011.

18. Upreti YR. Tobacco consumption among high school adolescents. Journal of Health Promotion. 2018;6:92–7.

19. Johnson RB, Christensen L. Educational research: Quantitative, qualitative and mixed approaches: SAGE; 2014.

20. Israel GD. Determining sample size: University of Florida; 1992/2013 [Available from: https://www.gjimt.ac.in/wp-content/uploads/2017/10/2_Glenn-D.-Israel_Determining-Sample-Size.pdf.

21. Ministry of Health Nepal, New ERA, ICF. Nepal Demographic and Health Survey 2016. 2017.

22. National Planning Commission. Fifteenth Planning. Kathmandu; 2020.

23. Bhaskar RK, Sah MN, Gaurav K, Bhaskar SC, Singh R, Yadav MK, et al. Prevalence and correlates of tobacco use among adolescents in the schools of Kalaiya, Nepal: A cross-sectional questionnaire based study Tobacco Induced Diseases. 2016;14:11.

24. Pradhan PMS, Niraula SR, Ghimire A, Singh SB, Pokharel PK. Tobacco use and associated factors among adolescent students in Dharan, Eastern Nepal: a cross-sectional questionnaire survey. BMJ open. 2013;3(2):e002123.

25. Ullah S, Sikander S, Abbasi MMJ, Rahim SA, Hayat B, Haq ZU, et al. Association between smoking and academic performance among under-graduate students of Pakistan, a cross-sectional study. 2019.

26. Joshi U, Pradhan M, Dahal S, Tyagi KK. Consumption of smokeless tobacco and areca nut among adolescents of bhaktpur, Nepal. Journal of Chitwan Medical College 2020;10(31):8–13.

27. Hussain A, Zaheer S, Shafique K. Individual, social and environmental determinants of smokeless tobacco and betel quid use amongst adolescents of Karachi: A school-based cross-sectional survey. BMC Public Health. 2017;17:913.

28. Kabir MA, Goh K-L. Determinants of tobacco use among students aged 13–15 years in Nepal and Sri Lanka: Results from the Global Youth Tobacco Survey, 2007. Health Education Journal. 2013;73(1):51–61.

29. Sreeramareddy CT, Kishore P, Paudel J, Menezes RG. Prevalence and correlates of tobacco use amongst junior collegiates in twin cities of western Nepal: A cross-sectional, questionnaire-based survey. BMC Public Health. 2008;8:97.

